# Epidemiological, virological and serological investigation into a SARS-CoV-2 outbreak (Alpha variant) in a primary school: a prospective longitudinal study

**DOI:** 10.1101/2021.10.26.21265509

**Authors:** Elsa Lorthe, Mathilde Bellon, Grégoire Michielin, Julie Berthelot, María-Eugenia Zaballa, Francesco Pennacchio, Meriem Bekliz, Florian Laubscher, Fatemeh Arefi, Javier Perez-Saez, Andrew S Azman, Arnaud G L’Huillier, Klara M Posfay-Barbe, Laurent Kaiser, Idris Guessous, Sebastian Maerkl, Isabella Eckerle, Silvia Stringhini, on behalf of the SEROCoV-Schools Study Group

## Abstract

We report a prospective epidemiological, virological and serological investigation of a SARS-CoV-2 outbreak in a primary school, as part of a longitudinal, prospective, primary school-based surveillance study. It involved repeated testing of pupils and teachers and household members of participants who tested positive, with rapid antigen tests and/or RT-PCR (Day 0-2 and Day 5-7), serologies on dried capillary blood samples (Day 0-2 and Day 30), contact tracing interviews and SARS-CoV-2 whole genome sequencing. This SARS-CoV-2 outbreak caused by the Alpha variant involved 20 children aged 4 to 6 years from 4 classes, 2 teachers and a total of 4 household members. Infection attack rates were between 11.8 and 62.0% among pupils from the 4 classes, 22.2% among teachers and 0% among non-teaching staff. Secondary attack rate among household members was 15.4%. Symptoms were reported by 63% of infected children, 100% of teachers and 50% of household members. All analysed sequences but one showed 100% identity. Serological tests detected 8 seroconversions unidentified by SARS-CoV-2 virological tests. This study confirmed child-to-child and child-to-adult transmission of the infection. Effective measures to limit transmission in schools have the potential to reduce the overall community circulation.

## Background

Children play an important role in the transmission of many respiratory viral diseases, including beta-coronaviruses and influenza virus, both within schools [1] and within households [2,3]. This has led most countries worldwide to implement school closures as an important component of severe acute respiratory syndrome coronavirus 2 (SARS-CoV-2) transmission mitigation policies from the very beginning of the pandemic [4].

Young children commonly have fewer and milder symptoms of SARS-CoV-2 infection than adults, with a high proportion of asymptomatic infections, and are less likely to experience severe COVID-19 [5]. However, epidemiological and biological data suggest that, when infected, children may transmit as much as adults, as children achieve viral loads comparable, or only minimally lower, to those of adults at the time of diagnosis [6–10].

At school, young children have many prolonged close contacts with peers and adults in indoor setting [11], usually do not wear masks, and in many countries they are not systematically tested when symptomatic, including in Switzerland [12]. These circumstances make children and schools a potential strong contributor of the overall community SARS-CoV-2 transmission [13]. Yet, almost two years into the pandemic, the extent to which young children are infected and transmit SARS-CoV-2 in school settings remains controversial [5,14], in particular with variants of concern. Evidence on transmission direction (adult-to-child, child-to-child, child-to-adult) is also lacking.

### Outbreak detection

This outbreak investigation is part of a longitudinal, prospective, observational surveillance study (SEROCoV-Schools), which aims to describe the transmission dynamics of SARS-CoV-2 infection within primary schools and early childhood education centres, and the risk of introduction of SARS-CoV-2 into the children’s households. The study started in March 2021.

Participants had a baseline assessment which included SARS-CoV-2 serology from a capillary blood test, an antigen rapid diagnostic test (RDT) from an oropharyngeal swab sample, and the completion of an online questionnaire. Then, a surveillance phase started, with weekly questionnaires and self-declarations (anytime outside of the weekly questionnaires) allowing participants to report COVID-19-like symptoms, contact with a positive case or the diagnosis of a SARS-CoV-2 infection. An outbreak investigation was triggered when a positive case was diagnosed from a positive real-time reverse transcription polymerase chain reaction (RT-PCR), which was the case in spring 2021.

In this study, we report the findings of a prospective epidemiological, virological and serological investigation of a SARS-CoV-2 outbreak in a primary school.

## Methods

### Study design and study population

The study population consisted of children and school staff in classes with a positive case as well as household members of the confirmed cases. School staff included teachers (including assistants) and non-teaching staff (administrative, cleaners, catering). Children absent from school for the 2 weeks preceding this outbreak were considered as non-exposed and not included in the analysis.

This prospective outbreak investigation involved repeated virological and serological testing of the participants. The date of diagnosis of the first positive case in a class was referred to as Day 0 for that class. RDT and/or RT-PCR were performed at Day 0-2 and Day 5-7. Serologies were performed at Day 0-2 and Day 30.

### Epidemiological investigation

#### Case definition

Cases were defined according to laboratory results: confirmed SARS-CoV-2-infected cases were those with positive SARS-CoV-2 RDT and/or RT-PCR results, and/or a seroconversion between D0-2 and D30 (from seronegative to seropositive according to the test-specific cut-off, unrelated to vaccination). Confirmed cases could be further classified as symptomatic or asymptomatic.

#### Other definitions

Infection attack rates (IAR) were defined as the proportion of all participating children and school staff with a positive RDT and/or RT-PCR, and/or a seroconversion. Household secondary attack rates (SAR) were defined as the proportion of household members who tested positive by RDT and/or RT-PCR result or who seroconverted, in households with one positive RDT and/or RT-PCR participant enrolled in the study. Adults were defined as individuals ≥18 years, whereas children were defined as individuals <18 years of age. Plausible directions of transmission were determined, when possible, on the basis of symptom onset and testing dates.

#### Contact tracing

Positive cases (or their parents in the case of children) were interviewed using a structured questionnaire investigating symptoms, contact with a positive person, school attendance, extracurricular activities, play dates/birthday parties, family/friend gatherings and best friends, in the 14 days prior to diagnosis.

### Laboratory investigation

#### Virological testing

SARS-CoV-2 testing was performed on oropharyngeal swabs taken by nurses or medical doctors of the study team. Children who were not attending school on the testing days were tested by their paediatrician or at a testing centre and communicated the results to our team. Depending on the visit, we performed RT-PCR or RDT tests. We used the Panbio COVID-19 Ag rapid test (Abbott) which has been validated in adults for use with oropharyngeal instead of nasopharyngeal swabs [15]. All oropharyngeal swab samples used for the RDT were tested a second time from the same swab by RT-PCR to confirm the result (in-house SARS-CoV-2 RT-qPCR or Cobas® SARS-CoV-2 Test, Cobas 6800, Roche, Switzerland). SARS-CoV-2 whole genome sequencing was performed for positive samples at the Health 2030 Genome Center (Geneva) using the Illumina COVIDSeq library preparation reagents following the protocol provided by the supplier.

#### Serological testing

We collected capillary blood on a Neoteryx Mitra® collection device, and tested for anti-Spike-SARS-CoV-2 IgG on a microfluidic nanoimmunoassay as described previously [16].

### Ethics

This study was approved by the ethics committee of the Canton of Geneva (Project ID 2020-02957). All parents and teachers were informed about the study and gave written consent while children gave verbal consent to participate.

### Role of the funding sources

The funders of the study had no role in the study design, data collection, data analysis, data interpretation, or writing of this manuscript.

## Results

### Outbreak description, epidemiological and serological investigation

The first COVID-19 case (triggering case: Teacher 1 [T1], Class 1) was diagnosed by RT-PCR in spring 2021, 3 days after the onset of symptoms. The second teacher in Class 1 (T2) had symptoms onset one day after T1 with an initially negative RT-PCR two days later followed by a positive RT-PCR two more days later (Figure 1).

**Figure 1:**
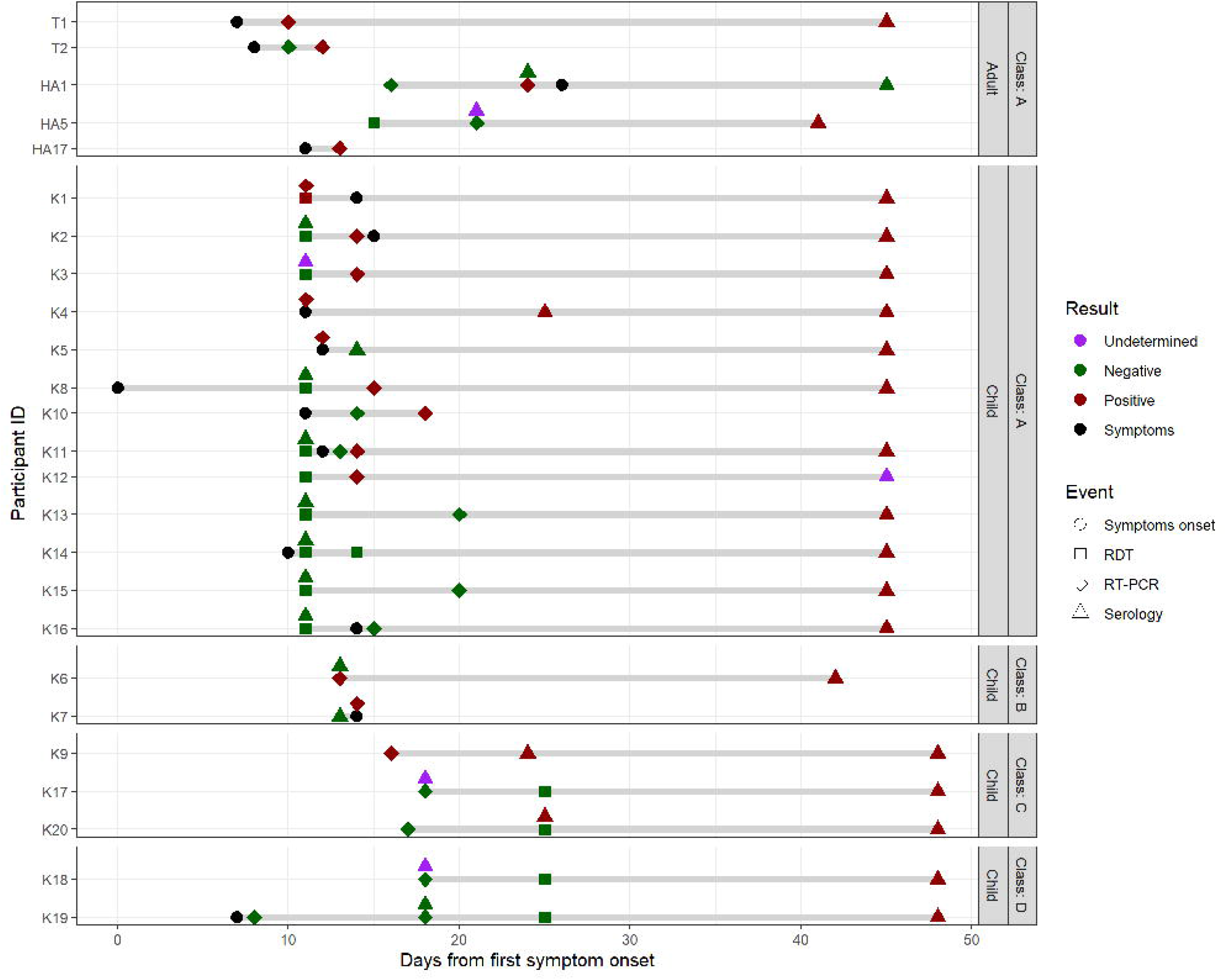
Timeline of symptoms onset, diagnosis and virological analyses among cases with a confirmed SARS-CoV-2 infection (positive RDT, RT-PCR or seroconversion) Legend: Apr: April, HAx: Adult member of household, HCx: Child member of household, Jun: June, Kx: Kid x, Tx: Teacher x, RDT: antigen rapid diagnostic test, RT-PCR: real-time reverse transcription polymerase chain reaction

All 21 children from Class 1, aged 4 years old, were tested the day after T1’s positive test. The IAR was 62% (13/21): 1 child had a positive RDT confirmed by a RT-PCR (Kid 1 [K1]); 5 children had a negative RDT, but subsequent RT-PCR testing on the same swab samples came back positive (K2, K3, K8, K11, K12); 2 children absent from school because symptomatic were tested by healthcare providers outside the study setting, both had a positive RT-PCR (K4, K5); 1 child had a positive RT-PCR at the second visit at D7 (K10); and 4 children seroconverted between the first and the last visit at D30 (K13, K14, K15, K16) despite negative swab virological tests at D1 and D7 (Table 1, Figures 1, 2 & 3).

**Table 1:** Overview of tests results, symptoms and cases among the 4 investigated classes, teachers and non-teaching staff

|  | Visit 1 (D0-2) |  | Visit 2 (D5-7)<sup>a</sup> |  | Seroconversion (D30)<sup>b</sup> | Symptoms<sup>c</sup> | Total cases<sup>d</sup> |
| --- | --- | --- | --- | --- | --- | --- | --- |
|  | RDT + | RT-PCR + | RDT + | RT-PCR + |  |  |  |
| Class A | 1/17 | 8<sup>e</sup>/21 | 0/1 | 1/16 | 4/11 | 10/13 | 13/21 |
| Class B | -/- | 2/17 | -/- | 0/15 | 0/12 | 1/2 | 2/19 |
| Class C | 0/1 | 1/17 | 0/14 | 0/2 | 2/16<sup>f</sup> | 0/3 | 3/18 |
| Class D | -/- | 0/15 | 0/12 | 0/1 | 2/10 | 1/2<sup>g</sup> | 2/15 |
| Teachers | 0/4 | 2/9 | 0/2 | 0/3 | 0/7 | 2/2 | 2/9 |
| Non-teaching staff | 0/2 | 0/10 | 0/4 | 0/2 | 0/10 | -/- | 0/13 |

<sup>a</sup> Tests were not repeated in participants with a SARS-CoV-2 infection diagnosed at Visit 1
<sup>b</sup> Among participants with negative swab tests both at Visit 1 and Visit 2, and negative serology at baseline and/or at Visit 1. Adults who were vaccinated and developed antibodies between visit 1 and visit 3 were not considered as related to the outbreak.
<sup>c</sup> Among participants with a SARS-CoV-2 positive test and/or a seroconversion
<sup>d</sup> A case was defined as a participant with a positive RDT and/or a positive RT-PCR and/or a seroconversion.
<sup>e</sup> Including confirmation of one positive RDT by a subsequent RT-PCR performed on the same day
<sup>f</sup> Including 1 seroconversion between March 26, 2021 and May 11, 2021
<sup>g</sup> No data on symptoms for one child

**Figure 2:**
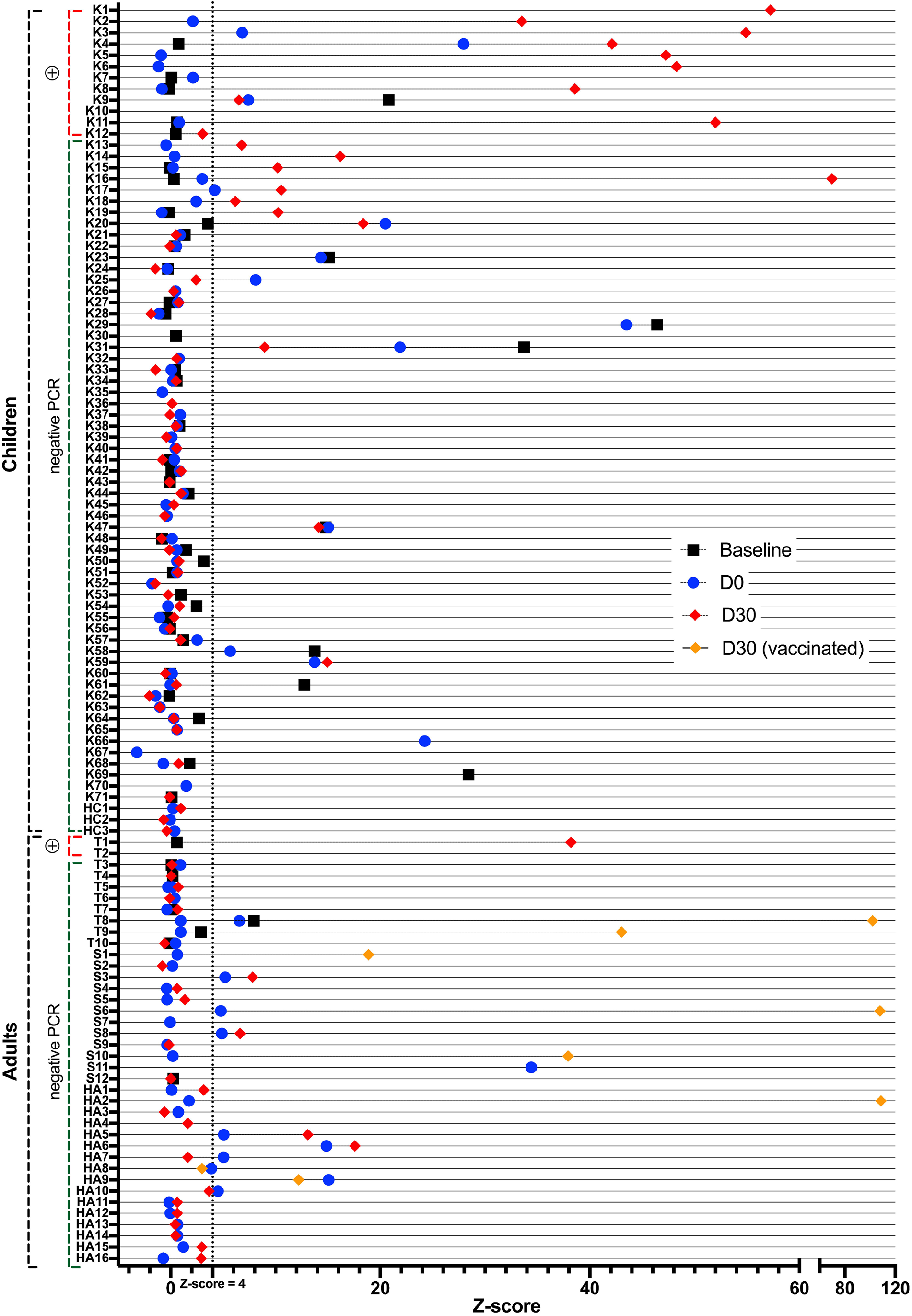
Results of serological tests at baseline, at D0-2 and D30 for all pupils, teachers, non-teaching staff and household members included in the outbreak investigation. Legend: This figure displays the results of the serological tests performed at baseline (black square), i.e. at the beginning of the study in March 2021, and during the outbreak investigation at Day 0-2 (blue circle) and Day 30 (red diamond). Adults who were vaccinated during the outbreak investigation are indicated by a yellow diamond. Household members who had no serological test are not represented. D: Day, HAx: Adult member of household, HCx: Child member of household, Kx: Kid x, Tx: Teacher x, PCR: real-time reverse transcription polymerase chain reaction

Children in another class from the same grade (Class 2) were identified as having close contacts with children of Class 1 during breaks and specific activities. Class 2 was therefore tested two days after the tests in Class 1. Of the 19 children aged 4 years old, 17 were tested by RT-PCR, of whom 2 (K6, K7) tested positive (IAR 11.8%), and 2 did not participate in the outbreak investigation (they both had only a negative RT-PCR performed externally at D8). No additional case was identified at subsequent visits. We also repeatedly tested 4 teachers and 2 non-teaching staff in contact with Classes 1 and 2, none of whom was diagnosed with a SARS-CoV-2 infection (Table 1).

The sibling of a positive case from Class 1, attending Class 3 in the same school, tested positive five days after his/her sibling (K9), triggering the testing protocol in Class 3. All 17 6-years old tested in Class 3 had a negative RT-PCR at D2, and 16/16 had a negative RDT at D9 (subsequently confirmed with a RT-PCR). Serological tests at D30 revealed 2 additional seroconversions (K17, K20, of whom one was not tested at D2 of the outbreak but seroconverted since baseline at the end of March 2021) (IAR 16.7%). In the other class from the same grade, Class 4, tested at the same time because of close and regular contacts with Class 3, 15/15 had a negative RT-PCR at D2, 13/13 had a negative RDT at D9, and 2 additional cases of seroconversion (K18, K19) were identified (IAR 13.3%); 2 children in the class did not take part in the study (Table 1, Figure 1). Among 3 teachers and 11 non-teaching staff, none tested positive.

Transmission occurred in 3 out of 10 investigated households of participants with a positive SARS-CoV-2 test, leading to a secondary attack rate among household members of 4/26 (15.4%). Three members of the same household refused to participate in this investigation, two of whom were vaccinated. Secondary attack rates were 1/2 (50.0%) with an adult index case, and 3/24 (12.5%) with a child index case. A teacher spent a few days at his/her parents’ place while having symptoms, his/her parent then developed symptoms and tested positive (adult household member [HA17]). The parent (HA5) of a positive child from Class 1 tested negative twice, though he/she seroconverted (Figure 1, Figure 2). He/she reported no contacts outside his/her household, strictly followed all recommendations, and was not vaccinated between the two blood draws. Finally, in the family of two children from Classes 1 and 3 who tested positive one after the other, a parent (HA1) who initially tested negative by RT-PCR, tested positive after quarantine with his/her children.

Among the 15 cases with a positive SARS-CoV-2 test, 3/3 (100%) adults and 9/12 (69%) children reported symptoms either before or after the positive test, and 3 children were asymptomatic. Among the 8 children who seroconverted without a positive RDT or RT-PCR test, 3 were symptomatic, 4 were asymptomatic, and one did not provide symptom information. Overall, 12/19 (63%) infected children reported symptoms. No severe form of COVID-19 (requiring hospitalization) was reported, and all cases recovered well.

### Contact tracing analysis

The two infected teachers live alone and reported no contact with a positive or symptomatic case in their private life or activities, during the 14 days prior to infection. Their presumed source of infection was school, as several children from Class 1 were coughing during the week of April 19. The two infected parents were likely infected by their children, as they reported no contact outside the household.

Three social activities outside the school setting occurred in the two weeks before the triggering case was diagnosed: the first one (8 days before the triggering case, outdoors) brought together 6 children from Class 1 of whom 5 subsequently tested positive and 3 kids from another school (not included in the study); the second one (5 days before the triggering case, both indoors and outdoors) gathered 4 children from Class 1 and Class 2 of whom 3 tested positive (the other one had antibodies at baseline and at Visit 1); the last one (2 days before the triggering case, outdoors) was attended by 2 kids from Class 1 and 2 kids from another school (not included in the study), 1 tested positive at the 2nd visit.

### Virological investigation

We conducted whole genome sequencing on samples from the 15 participants who tested positive for SARS-CoV-2. Coverage of three isolate genomes (HA1, K9 and K10) were insufficient for any comparisons due to low viral load in the specimen (32%, 20% and 16% coverage only, respectively). The other sequenced genomes belonged to the Alpha variant. The virus sequence of K12 differed from the others in one region covered. Here, in position 15824-15827 a deletion and one addition that restores the reading frame was observed, resulting in a total of 4 nucleotides difference compared to the other sequences. All 9 sequences with >99% coverage shared 100% identity between genomes in comparison to the reference sequence (Figure 3). Virus specimens that could only be partially sequenced retrieved the same sequence without additional mutations in the regions covered.

**Figure 3:**
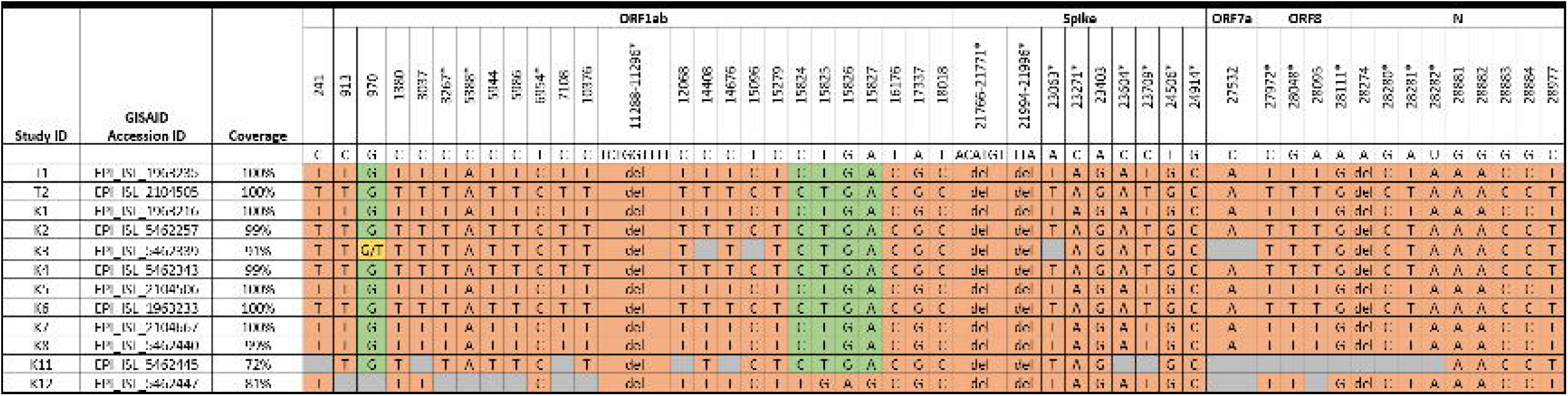
Virological analysis of positive cases by SARS-CoV-2 full genome sequencing. ID: Identifier, GISAID: Global Initiative on Sharing Avian Influenza Data, Kx: Kid x, Tx: Teacher x Legend: Mutations in comparison to the reference sequence (NC_045512) are highlighted in orange. Green fields indicate no mutation; grey fields indicate insufficient genome coverage; and yellow fields indicate mixed viral population of the two nucleotides given. Numbers indicate nucleotide positions; asterisks (^*^) mark lineage-defining mutations for the Alpha variant.

## Preventive measures and outbreak control measures

During the period of this outbreak investigation, non-pharmaceutical public health measures in Geneva were gradually relaxed, with restaurants and bars opening their outdoor spaces and entertainment venues opening their indoor spaces.

Mitigation of the coronavirus disease (COVID-19) pandemic in Switzerland included school closures during the first wave from March to May 2020. Thereafter, priority was given to keeping schools open with several types of preventive measures in place which varied widely across institutions. In the investigated school, measures in place during the outbreak included checking children’s temperature every morning, sending children home if they had fever or sickness beyond very mild symptoms, restricted access for parents and requiring all adults to wear facemasks.

During the outbreak, classes 1 and 2 were placed in quarantine by local health authorities for 10 days, starting 4 days after the triggering case. Classes 3 and 4 were not placed in quarantine.

## Discussion

This is the first investigation of a SARS-CoV-2 outbreak caused by the Alpha variant in a primary school. It involved 20 children from 4 classes, 2 teachers and 4 household members (of whom one was also attending one of the participating classes). The index case could not be formally identified, but it is likely that this outbreak was only identified a week after it started when a teacher tested positive and triggered the investigation. Of note, children in this age group were not routinely tested by the official testing recommendations in Switzerland when symptomatic [12]. This prospective classroom-based study provides evidence of SARS-CoV-2 circulation among young children, school teachers, and introduction into households.

Since at least 9 positive cases of this outbreak shared viruses with identical sequence, we conclude that they are part of the same cluster. This could have been either by simultaneous infection through the same source, or transmission chains between affected individuals. One divergent virus sequence was found in one of the positive children, which could be either a *de novo* mutation occurring during the outbreak, or constitute independent introductions into the school, with one leading to this cluster. It cannot be ruled out that, by coincidence, several infection events with viruses sharing the same sequence were introduced from the community independently into the school. However, this hypothesis seems less likely given the number of cases involved, the epidemiological link and time frame, and the limited period of potentially other exposures before the quarantine decision. Overall community circulation at the cantonal level was also relatively low at the time (weekly incidence: about 200 cases/100,000 inhabitants), while the Alpha variant was causing 92% of new SARS-CoV-2 infections in Geneva [17].

Other main insights are as follows. First, viral circulation of the Alpha variant in young children aged 4-6 years old was high, with a majority of unspecific and mild symptomatic infections, which might explain relatively high secondary attack rates [13]. The observed cluster probably started in one class and spread to two other classes, most likely through direct child-to-child contacts and interactions as per our epidemiological investigation, although there could be non-sampled child or adult intermediaries. We identified two seroconversions in a fourth class, but with no identification of SARS-CoV-2 virus, thereby limiting the conclusions on transmission linked to the other classes.

Second, child-to-parent transmission occurred in two different households, and child-to-teacher transmission is probable, which supports previous findings [18,19]. Of note, vaccination was opened to all persons aged 45 years or older from April 12, and to all people in the 16-44 age group from May 19. Therefore, only a minority of adults (1 household member and 1 non-teaching staff) were vaccinated at the beginning of the outbreak. Child-to-adult transmission seems to depend on the duration of contacts, as no infection was identified among non-teaching staff who spent only limited periods of time (meals) with children. This is contrary to previous findings [20], and may be explained by the young age of our participants and their behaviour involving physical proximity with their teachers [21].

Third, social activities outside the school could have contributed to the spread of the infection, as previously reported [22]. However, they could also reflect the bonding between children and a closer contact at school, thus facilitating transmission.

Fourth, we evidenced low sensitivity of RDTs with an oropharyngeal specimen for identifying both symptomatic and asymptomatic infected children, which confirms previous results showing that only children with high viral load are identified by such tests [23,24]. We conclude that RDTs with an oropharyngeal specimen are not the most appropriate for surveillance and/or outbreak investigation purposes. Analysis of joint RT-PCR/serological data shows that there is a substantial under-detection of infections in young children, even with RT-PCR testing, although the optimal time point for viral testing might have been missed in the fixed testing scheme of this study. Under-detection of acutely infected children might be an explanation for the discrepancy between official numbers of infected children [25], and studies on virus prevalence and seroprevalence in school settings [26–30].

### Strengths and limitations

Few large SARS-CoV-2 outbreaks in young children in school settings have been documented so far [31,32], even fewer involved an investigation of variants of concern. As part of an ongoing prospective study, this investigation started less than 24 hours after the first case was laboratory-confirmed and involved the use of three complementary approaches. We followed up and repeatedly tested all contacts within four classes regardless of symptoms. Repeated serological tests proved useful to retrieve seroconversions following asymptomatic or undiagnosed infections. However, we relied on a limited number of cases. Not all children and adults were tested, which could lead to underestimating IAR and SAR. We might have missed infections among adults who were vaccinated between D0-2 and D30, as we could not distinguish between antibodies due to vaccination and those due to infection. Also, we could not test the household members of cases only detected by seroconversion at D30 with no positive RT-PCR/RDT, leading to a potential underestimation of secondary attack rates. The study was performed before the circulation of the more infectious Delta variant; estimates are therefore likely to be underestimated in a context of Delta dominance [33].

## Conclusion

This prospective school-based study provides evidence of SARS-CoV-2 transmission among young children and school teachers and introduction into households. Epidemiological investigation confirmed child-to-child and child-to-adult direction of transmission of the infection. Children may be a significant source of extra-household infections and have the potential to play a role in community transmission, potentially even more so with the more contagious Delta variant. With most of the adult and adolescent population vaccinated, children could act as disease reservoirs. Effective strategies are needed to limit transmission in school settings and vaccination of school staff, and children when available, should be encouraged.

## Data Availability

Our data are accessible to researchers upon reasonable request for data sharing to Silvia Stringhini.

## Conflict of interest statement

The authors have no conflict of interest in relation with this article.

## Funding

The SEROCoV-Schools study was supported by the Federal Office of Public Health, the Private Foundation of the Geneva University Hospitals, the Fondation des Grangettes, the Center for Emerging Viral Diseases, and a SNF NRP (National Research Program) 78 Covid-19 Grant 198412 (to S.J.M., I.E.).

## Contributors

SS, IE, ALH, KPB, AA, EL, MBel and IG contributed to the conceptualisation of the study. EL, JB, MEZ, FP, SS and MBel participated in the project management and implementation. EL performed all contact-tracing interviews. GM, FA and SM analysed all serologies. MBel, MBek, FL, LK and IE performed and interpreted all virological analyses. GM, JPS, MBel and IE did the figures and visualisations. EL and MBel verified the data. All authors had full access to all the data in the study, participated in the interpretation of the results, and accept responsibility to submit for publication. EL wrote the first draft, and all other authors reviewed the manuscript. The SEROCoV-Schools Study Group contributed to the project implementation.

## Acknowledgements

We are very grateful to the children and teachers participating in the SEROCoV-Schools study and their families. We would like to thank the school staff for their responsiveness and support during the outbreak investigation. The authors also would like to thank the members of the Unit of Population Epidemiology for their daily support in all the tasks required by the SEROCoV-School study.

## SEROCoV-Schools Study Group

Elsa Lorthe, Julie Berthelot, Maria-Eugenia Zaballa, Hélène Baysson, Andrea Jutta Loizeau, Ania Wisniak, Stéphanie Testini, Khadija Samir, Natalie Francioli, Severine Harnal, Javier Perez-Saez, Nick Pullen, Francesco Pennacchio, Julien Lamour, Gaëlle Bryand-Rumley, Claire Semaani, Viviane Richard, Roxane Dumont, Prune Collombet, Natacha Noël, Patrick Bleich, Nacira El Merjani, Caroline Pugin, Jessica Rizzo, Marion Frangville, Antoine Bal, Fanny-Blanche Lombard, Zo Francia Randrianandrasana, Oumar Aly Ba, Chantal Martinez, Paola D’Ippolito, Camille Tible, Viola Bucolli, Livia Boehm, Adrien Jos Rastello, Lucie Ménard, Lison Beigbeder, Fréderic Rinaldi, Alain Cudet, Alexandre Moulin, Andrew S Azman, Arnaud G L’Huillier, Klara M Posfay-Barbe, Idris Guessous, Silvia Stringhini, Grégoire Michielin, Sebastian Maerkl, Fatemeh Arefi, Mathilde Bellon, Isabella Eckerle, Laurent Kaiser, Benjamin Meyer, Meriem Bekliz, Florian Laubscher, Francisco Perez Rodriguez, Pascale Sattonnet-Roche, Catia Alvarez, Kenneth Adea, Manel Essaidi-Laziosi, Gil Barbosa Monteiro

## Notes

### Competing Interest Statement

The authors have declared no competing interest.

### Author Declarations

This study was approved by the ethics committee of the Canton of Geneva (Project ID 2020-02957).

### Summary of Updates

Precise dates and places were removed from the manuscript.

## References

1. Marchbanks TL, Bhattarai A, Fagan RP, Ostroff S, Sodha SV, Moll ME, et al. An Outbreak of 2009 Pandemic Influenza A (H1N1) Virus Infection in an Elementary School in Pennsylvania. Clin Infect Dis. 2011;52(Suppl 1):S154–60.

2. Cauchemez S, Donnelly CA, Reed C, Ghani AC, Fraser C, Kent CK, et al. Household Transmission of 2009 Pandemic Influenza A (H1N1) Virus in the United States. New England Journal of Medicine. 2009;361(27):2619–27.

3. Monto AS, DeJonge PM, Callear AP, Bazzi LA, Capriola SB, Malosh RE, et al. Coronavirus Occurrence and Transmission Over 8 Years in the HIVE Cohort of Households in Michigan. J Infect Dis. 2020;222(1):9–16.

4. Viner RM, Russell SJ, Croker H, Packer J, Ward J, Stansfield C, et al. School closure and management practices during coronavirus outbreaks including COVID-19: a rapid systematic review. Lancet Child Adolesc Health. 2020;4(5):397–404.

5. Wiedenmann M, Goutaki M, Keiser O, Stringhini S, Tanner M, Low N. The role of children and adolescents in the SARS-CoV-2 pandemic: a rapid review. Swiss Medical Weekly. 2021;151:w30058.

6. Zhu Y, Bloxham CJ, Hulme KD, Sinclair JE, Tong ZWM, Steele LE, et al. A Meta-analysis on the Role of Children in Severe Acute Respiratory Syndrome Coronavirus 2 in Household Transmission Clusters. Clinical Infectious Diseases. 2021;72(12):e1146–53.

7. Jones TC, Biele G, Mühlemann B, Veith T, Schneider J, Beheim-Schwarzbach J, et al. Estimating infectiousness throughout SARS-CoV-2 infection course. Science. 2021;373(6551):eabi5273.

8. Baggio S, L’Huillier AG, Yerly S, Bellon M, Wagner N, Rohr M, et al. SARS-CoV-2 viral load in the upper respiratory tract of children and adults with early acute COVID-19. Clin Infect Dis. 2020;ciaa1157.

9. L’Huillier AG, Torriani G, Pigny F, Kaiser L, Eckerle I. Culture-Competent SARS-CoV-2 in Nasopharynx of Symptomatic Neonates, Children, and Adolescents. Emerg Infect Dis. 2020;26(10):2494–7.

10. Bellon M, Baggio S, Bausch FJ, Spechbach H, Salamun J, Genecand C, et al. SARS-CoV-2 viral load kinetics in symptomatic children, adolescents and adults. Clin Infect Dis. 2021;ciab396.

11. Stehlé J, Voirin N, Barrat A, Cattuto C, Isella L, Pinton J-F, et al. High-Resolution Measurements of Face-to-Face Contact Patterns in a Primary School. PLoS One. 2011;6(8):e23176.

12. BAG. COVID-19 - Recommandations sur la procédure à suivre pour les enfants symptomatiques de moins de 6 ans ainsi que les autres personnes fréquentant les écoles et les structures d’accueil parascolaire/extrafamilial et indications de test chez les enfants de moins de 6 ans [Internet]. 2021. Disponible sur: https://cdn.paediatrieschweiz.ch/production/uploads/2021/03/2021.03.23-Indications-de-test-chez-les-enfants-symptomatiques-de-moins-de-6-ans.pdf

13. Cevik M, Marcus JL, Buckee C, Smith TC. Severe Acute Respiratory Syndrome Coronavirus 2 (SARS-CoV-2) Transmission Dynamics Should Inform Policy. Clinical Infectious Diseases. 2021;73(Supplement_2):S170-6.

14. Yung CF, Kam K, Nadua KD, Chong CY, Tan NWH, Li J, et al. Novel Coronavirus 2019 Transmission Risk in Educational Settings. Clinical Infectious Diseases. 2021;72(6):1055–8.

15. Nsoga MTN, Kronig I, Rodriguez FJP, Sattonnet-Roche P, Silva DD, Helbling J, et al. Diagnostic accuracy of Panbio rapid antigen tests on oropharyngeal swabs for detection of SARS-CoV-2. PLOS ONE. 2021;16(6):e0253321.

16. Swank Z, Michielin G, Yip HM, Cohen P, Andrey DO, Vuilleumier N, et al. A high-throughput microfluidic nanoimmunoassay for detecting anti–SARS-CoV-2 antibodies in serum or ultralow-volume blood samples. Proc Natl Acad Sci USA. 2021 May 4;118(18):e2025289118

17. Althaus CL, Baggio S, Reichmuth ML, Hodcroft EB, Riou J, Neher RA, et al. A tale of two variants: Spread of SARS-CoV-2 variants Alpha in Geneva, Switzerland, and Beta in South Africa [Internet]. 2021 june. doi: https://doi.org/10.1101/2021.06.10.21258468

18. Ismail SA, Saliba V, Bernal JL, Ramsay ME, Ladhani SN. SARS-CoV-2 infection and transmission in educational settings: a prospective, cross-sectional analysis of infection clusters and outbreaks in England. The Lancet Infectious Diseases. 2021;21(3):344–53.

19. Meuris C, Kremer C, Geerinck A, Locquet M, Bruyère O, Defêche J, et al. Transmission of SARS-CoV-2 After COVID-19 Screening and Mitigation Measures for Primary School Children Attending School in Liège, Belgium. JAMA Network Open. 2021;4(10):e2128757.

20. Ladhani SN, Baawuah F, Beckmann J, Okike IO, Ahmad S, Garstang J, et al. SARS-CoV-2 infection and transmission in primary schools in England in June–December, 2020 (sKIDs): an active, prospective surveillance study. The Lancet Child & Adolescent Health. 2021;5(6):417–27.

21. Paul LA, Daneman N, Schwartz KL, Science M, Brown KA, Whelan M, et al. Association of Age and Pediatric Household Transmission of SARS-CoV-2 Infection. JAMA Pediatr. 2021;175(11):1151–1158.

22. Ghinai I, Woods S, Ritger KA, McPherson TD, Black SR, Sparrow L, et al. Community Transmission of SARS-CoV-2 at Two Family Gatherings — Chicago, Illinois, February– March 2020. MMWR Morb Mortal Wkly Rep. 2020;69(15):446–50.

23. Kriemler S, Ulyte A, Ammann P, Peralta GP, Berger C, Puhan MA, et al. Surveillance of Acute SARS-CoV-2 Infections in School Children and Point-Prevalence During a Time of High Community Transmission in Switzerland. Front Pediatr. 2021;9:645577.

24. L’Huillier AG, Lacour M, Sadiku D, Gadiri MA, De Siebenthal L, Schibler M, et al. Diagnostic Accuracy of SARS-CoV-2 Rapid Antigen Detection Testing in Symptomatic and Asymptomatic Children in the Clinical Setting. J Clin Microbiol. 59(9):e00991–21.

25. Posfay-Barbe KM, Andrey DO, Virzi J, Cohen P, Pigny F, Goncalves AR, et al. Prevalence of IgG against SARS-CoV-2 and evaluation of a rapid MEDsan IgG test in children seeking medical care. Clin Infect Dis. 2020;ciaa1702.

26. Villani A, Coltella L, Ranno S, Bianchi di Castelbianco F, Murru PM, Sonnino R, et al. School in Italy: a safe place for children and adolescents. Ital J Pediatr. 2021;47:23.

27. Varma JK, Thamkittikasem J, Whittemore K, Alexander M, Stephens DH, Arslanian K, et al. COVID-19 Infections Among Students and Staff in New York City Public Schools. Pediatrics (2021) 147 (5): e2021050605.

28. Hommes F, van Loon W, Thielecke M, Abramovich I, Lieber S, Hammerich R, et al. SARS-CoV-2 Infection, Risk Perception, Behaviour, and Preventive Measures at Schools in Berlin, Germany, during the Early Post-Lockdown Phase: A Cross-Sectional Study. Int J Environ Res Public Health. 2021;18(5):2739.

29. Fontanet A, Tondeur L, Grant R, Temmam S, Madec Y, Bigot T, et al. SARS-CoV-2 infection in schools in a northern French city: a retrospective serological cohort study in an area of high transmission, France, January to April 2020. Eurosurveillance. 2021;26(15):2001695.

30. Dattner I, Goldberg Y, Katriel G, Yaari R, Gal N, Miron Y, et al. The role of children in the spread of COVID-19: Using household data from Bnei Brak, Israel, to estimate the relative susceptibility and infectivity of children. PLoS Comput Biol. 2021;17(2):e1008559.

31. Stein-Zamir C, Abramson N, Shoob H, Libal E, Bitan M, Cardash T, et al. A large COVID-19 outbreak in a high school 10 days after schools’ reopening, Israel, May 2020. Eurosurveillance. 2020;25(29):2001352.

32. Torres JP, Piñera C, De La Maza V, Lagomarcino AJ, Simian D, Torres B, et al. SARS-CoV-2 antibody prevalence in blood in a large school community subject to a Covid-19 outbreak: a cross-sectional study. Clin Infect Dis. 2020;ciaa955.

33. Li B, Deng A, Li K, Hu Y, Li Z, Xiong Q, et al. Viral infection and transmission in a large, well-traced outbreak caused by the SARS-CoV-2 Delta variant [Internet]. 2021 july. doi: https://doi.org/10.1101/2021.07.07.21260122

